# Association of age at menopause and risk of depression during the perimenopause and postmenopause

**DOI:** 10.64898/2026.08.13.26360384

**Authors:** Rochelle Knight, Carol Joinson, Abigail Fraser, Kimberly Burrows, Ana Gonçalves Soares

## Abstract

**Importance:** The menopausal transition has been associated with an increased risk of depression, although findings are inconsistent. While most research has focused on menopausal stage, some studies suggest that later age at menopause may be associated with lower depression risk.

**Objective:** To examine the association between age at menopause and depression risk during perimenopause and early postmenopause using multivariable regression and genetic approaches.

**Design:** Prospective cohort study using data from the mothers of the Avon Longitudinal Study of Parents and Children (ALSPAC), a UK birth cohort that recruited pregnant women in 1991-1992.

**Setting:** UK community-based cohort study.

**Participants:** Up to 3,307 women with repeated measures of depressive symptoms across the perimenopausal and postmenopausal periods and data on observed or genetically predicted age at menopause.

**Exposure:** Observed age at menopause, a polygenic risk score (PRS) for age at menopause, and genetically predicted age at menopause.

**Main Outcome(s) and Measure(s):** Depressive symptoms during the perimenopausal and early postmenopausal periods were assessed using the Edinburgh Postnatal Depression Scale (EPDS), with depression defined as a score ≥ 13.

**Results:** Effect estimates across multivariable regression and genetic analyses were small and directionally consistent with lower odds of depression with older age at menopause, although most confidence intervals included the null. In analyses using observed age at menopause, there was little evidence of an association with depression during perimenopause (Odds ratio (OR) per year increase in age at menopause 0.98, 95%CI 0.89–1.08) or postmenopause (OR 1.00, 95%CI 0.89–1.13). Results were similar when using a PRS as a genetic proxy for age at menopause during perimenopause (OR per standard deviation (SD) increase in PRS 0.98, 95%CI 0.89–1.09) but suggested lower odds of depression during postmenopause (OR 0.92, 95%CI 0.86–0.99). Mendelian randomization analyses did not support a causal effect (OR per year increase 1.00, 95%CI 0.89–1.13 for perimenopause, and OR 0.97, 95%CI 0.86–1.09 for postmenopause).

**Conclusions and Relevance:** Age at menopause is unlikely to be a major driver of midlife depression risk. However, consistent effect directions across approaches suggest a small association may exist, but further research in larger samples is needed to confirm this.

**Key points:** *Question:* Is age at menopause associated with risk of depression during perimenopause and early postmenopause?

*Findings:* In this prospective cohort study of up to 3,307 women, effect estimates across multivariable regression and genetic analyses were directionally consistent suggesting lower depression risk with later age at menopause, although estimates were small and evidence was limited.

*Meaning:* Our findings suggest that age at menopause is unlikely to be a major contributor to depression risk during the menopausal transition.

## Introduction

Women experience major depression at approximately twice the rate of men^1–3^, with the greatest disparity during the reproductive years^4^ and attenuating in later life^5^. Evidence suggest that the menopausal transition may be associated with an increased risk of depression,^6–10^ but findings are inconsistent^7,8^. A recent meta-analysis of longitudinal studies reported higher depression risk during perimenopause compared with premenopause (odds ratio [OR] 1.40, 95% confidence interval [95%CI] 1.21, 1.61), with no clear increase in postmenopause (OR 1.18, 95%CI 0.74, 1.86).^9^

Variation in the timing of menopause may also be important. The mean age at natural menopause is approximately 50.5 years, although this varies across populations,^11^ and it is influenced by socioeconomic, behavioural and reproductive factors.^12^ Earlier menopause has been associated with adverse health outcomes, including cardiovascular disease^13^ and all-cause mortality^13^. There is also evidence that a later age at menopause is associated with a decreased risk of depression^14,15^.

However, much of the evidence is limited by cross-sectional designs^14^, retrospective recall of menopausal age^13,14^, and a focus on postmenopausal depression^14^, with less attention to the perimenopausal period.

Triangulation of evidence across complementary analytic approaches may help strengthen causal inference, as different methods are subject to distinct sources of bias and rely on different assumptions.^16^ Genetic approaches may be particularly useful for addressing limitations of traditional observational studies, including residual confounding and reserve causation. Polygenic risk scores (PRS) provide an estimate of genetic liability to traits such as earlier or later age at natural menopause^17^. Because genetic variants are randomly allocated at conception, PRS analyses are less susceptible to reverse causation and environmental confounding. However, PRS-based associations reflect genetic liability rather than a causal effect and may be influenced by horizontal pleiotropy or shared biological pathways.^17,18^ Mendelian randomisation (MR) extends this framework by using genetic variants as instrumental variables to estimate the causal effect of age at menopause on depression.^17–20^ Unlike using a PRS, MR explicitly targets a causal estimate, although validity depends on core assumptions.

In this study, we used data from a large prospective community-based cohort to:

1. Examine the prospective association between observed age at natural menopause and depression during the perimenopausal and early postmenopausal periods.
2. Investigate menopausal timing using genetic approaches by examining associations between a PRS for age at menopause and depression, and applying one-sample MR to estimate the potential causal effect of menopausal timing on depression.

## Methods

### Participants

This study uses data from the mothers cohort of the Avon Longitudinal Study of Parents and Children (ALSPAC). The study invited pregnant women residing in Avon, UK with an expected delivery date between 1^st^ April 1991 and 31^st^ December 1992 to take part. The initial ALSPAC sample enrolled 14,541 pregnancies from 14,203 unique mothers, resulting in 13,988 children who were alive at 1 year of age. As a result of additional phases of recruitment, a further 630 joined the study bringing the total to 14,833 unique mothers (known as Generation 0 or G0). Since enrolment, participants and their families have been followed through questionnaires, research clinic assessments, and data linkage. Since 2014 study data have been collected and managed using REDCap (Research Electronic Data Capture), a secure, web-based platform hosted at the University of Bristol^21^. REDCap is specifically designed to support data capture and management in research studies.

The cohort’s recruitment, representativeness, and profile have been extensively described elsewhere^22–25^. A fully searchable data dictionary and variable search tool are available on the ALSPAC website (http://www.bristol.ac.uk/alspac/researchers/our-data).

### Measures

#### Depression outcome

Depressive symptoms were assessed using the 10-item Edinburgh Postnatal Depression Scale (EPDS)^26^. While originally developed to assess perinatal depression, the EPDS has since been widely used to evaluate depressive symptoms in adults beyond the perinatal period^27–30^. A score of 13 or higher is commonly used to indicate probable depression^26,31^.

Since enrolment, the EPDS was administered at 15 timepoints spanning over 30 years. Our analyses used 11 timepoints outside the perinatal period (pregnancy and the first year postpartum).

Information on current antidepressant use was collected at seven timepoints.

Probable depression (hereafter called depression) was defined as either (i) an EPDS score ≥ 13 or (ii) reported current use of antidepressant medication. At timepoints where both measures were available, depression was defined as meeting either criterion.

Details on the variables used to derive the depression outcome are presented in eTables 1-2.

#### Age at Menopause and reproductive stage

Age at natural menopause was estimated based on self-reported menstrual bleeding patterns^32^. The final menstrual period (FMP) was defined as the date of the last menstrual period followed by at least 12 months of amenorrhea. Age at natural menopause (hereafter referred to as age at menopause) was defined as the age at FMP.

In addition, all women who provided menstrual cycle data – regardless of whether an age at menopause could be estimated – were classified into reproductive stage (reproductive, perimenopause and postmenopause) at each depression assessment according to the Stages of Reproductive Ageing Workshop + 10 (STRAW+10)^33^ criteria.

Reproductive stage was assigned at each depression assessment and used to determine depression outcomes as occurring during the perimenopausal and postmenopausal period. Staging did not require estimation of age at menopause, allowing inclusion of women without a known age at menopause in analyses that did not require the observed exposure (e.g. when using a PRS for the exposure).

Further details on how reproductive stage was assigned are provided in Supplementary Methods.

#### Confounders

Confounders were selected a priori based on prior literature. All confounders were based on self-report and included: age, ethnicity, social class, age at menarche, educational attainment, material hardship, social support, smoking status, body mass index (BMI) and alcohol intake. Further details on the specific questionnaire items and variable coding are provided in eTable 3 and Supplementary Methods. Given some studies report a possible link between history of depression and age at menopause^34,35^, we additionally adjusted for a history of depression and history of antidepressant use prior to the perimenopausal period in sensitivity analyses.

Ethnicity, social class, age at menarche, and educational attainment were treated as baseline confounders. Material hardship, social support, smoking status, BMI, and alcohol intake were treated as time-varying confounders.

### Statistical analysis

Three complementary analytic approaches were used to examine the association between age at menopause and depression during the perimenopausal and postmenopausal periods: (i) multivariable regression using observed age at menopause; (ii) multivariable regression using a PRS for age at menopause (PRS_ANM_) as a genetic proxy; and (iii) one-sample Mendelian randomisation using PRS_ANM_ as an instrumental variable.

These genetic approaches provide complementary strengths. Conventional multivariable regression estimates associations on the exposure scale (e.g. OR per year increase in age at menopause) but may be susceptible to residual and unmeasured confounding. Using a PRS as the exposure in multivariable regression reduces susceptibility to environmental confounding, as genetic variants are fixed at conception and are generally not be associated with environmental or behavioural factors at the population level. ^17,18^ This approach also increases statistical power by allowing inclusion of participants with genetic data but missing phenotypic menopause data, although estimates are less directly interpretable (e.g. per standard deviation (SD) increase in PRS) and do not provide causal inference.

One-sample MR extends this framework by using genetic variants as instrumental variables to strengthen causal inference^17–20^ and provide estimates on the exposure scale. However, causal interpretation depends on key core assumptions.^36^ In contrast to MR, PRSs are optimised for prediction rather than causal inference and may include variants with pleiotropic effects that violate instrumental variable assumptions. MR analyses also typically have lower statistical power because both genetic and phenotypic data are required. Each approach is described in further detail below.

Analyses included women with at least one measure of depression during the perimenopausal or postmenopausal period. Observations following hysterectomy, oophorectomy, or more than 12 months of amenorrhea attributed to surgery were excluded. For analyses requiring estimated age at menopause, women with an estimated age at menopause younger than 40 years (likely reflecting premature ovarian insufficiency) were excluded (N = 69). For analyses using observed age at menopause, postmenopausal observations were restricted to 8 years postmenopause in line with the STRAW+10 definition of early postmenopause. Sample derivation is shown in Figure 1.

**Figure 1.**
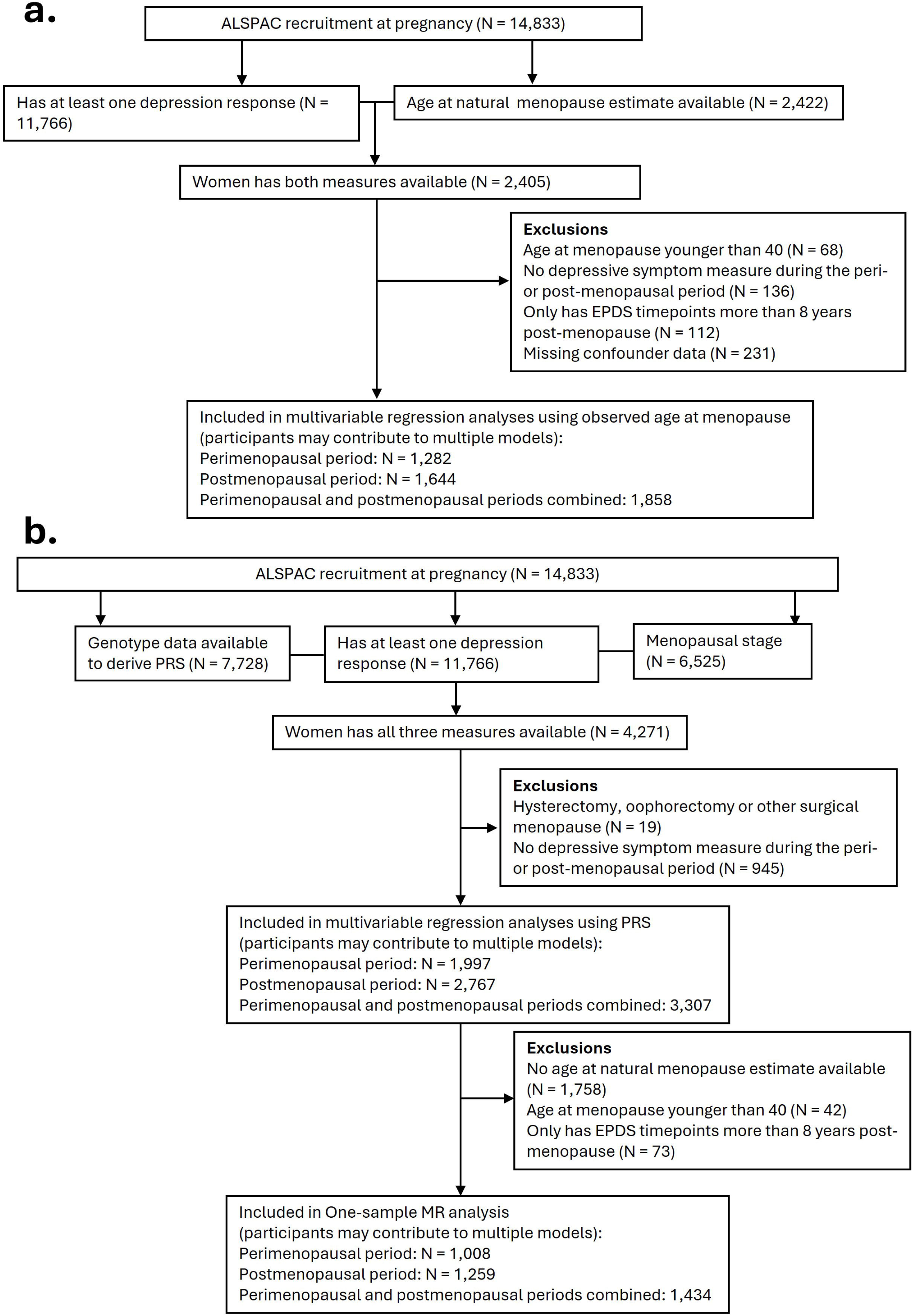
Flowchart of participants included in the analyses. (A) shows the flow for the multivariable regression. (B) shows the flow for the PRS regression and One-sample Mendelian Randomisation.

Associations were analysed separately for the perimenopausal and postmenopausal periods, with secondary analyses combining these stages. Baseline characteristics were compared between analytic samples.

#### Multivariable regression analysis using observed age at menopause

The association between observed age at menopause and depression during the perimenopausal and postmenopausal periods was assessed using random-effects binomial logistic regression to account for repeated depression measures within individuals. Models included a participant-specific random intercept and were adjusted for the confounders defined above. Odds ratios (ORs) of depression were estimated per one-year increase in age at menopause.

#### Multivariable regression analysis using genetic proxy for age at menopause (PRS_ANM_)

A PRS for age at menopause (PRS_ANM_) was constructed for all participants with genotype data. Details on how ALPSAC mothers were genotyped are described in Supplementary methods. Genetic data was available for 7,728 unrelated mothers. After restricting to those with a perimenopausal or postmenopausal depression measure the final sample was 3,307 (Figure 1).

Summary statistics from a genome-wide association study (GWAS) meta-analysis of age at natural menopause^37^ was used to identify single nucleotide polymorphisms (SNPs) associated with age at natural menopause.

PRS_ANM_ were generated using PRSice-2^38^, with linkage disequilibrium (LD) clumping applied (r^2^ < 0.05 within a 1-Mb window). Scores were weighted by GWAS effect estimate size and standardised to a mean of 0 and standard deviation of 1. Scores were constructed across a range of GWAS *p*-value thresholds for SNP inclusion (5x10^-8^ to 1; eTable 4). The best-fit PRS_ANM_ was determined by evaluating its association with the age at menopause phenotype derived in ALSPAC (Supplementary Methods).

The association between the best-fit PRS_ANM_ and depression during the perimenopausal and postmenopausal periods was assessed using random-effects binomial logistic regression. Models included a participant-specific random intercept and were adjusted for age and the first ten principal components (PCs) of ancestry. ORs were estimated per standard deviation (SD) increase in PRS_ANM_.

#### One-sample Mendelian randomisation

One-sample MR was used to estimate the causal effect of age at menopause on the risk of depression during the perimenopausal and postmenopausal periods. Depression was defined as binary indicators of any occurrence of depression within the perimenopausal or postmenopausal period, with outcomes defined separately for each stage.

One sample MR was implemented using two-stage residual inclusion (TSRI) via the tsri() function from the *OneSampleMR* R package, adjusting for the first 10 principal components of ancestry. Effects estimates are presented as ORs per one-year increase in age at menopause. Further details are provided in ematerials.

### Sensitivity analyses

To assess the impact of restricting postmenopausal observations to within 8 years of menopause, we repeated the multivariable regression using observed age at menopause and one-sample MR analyses without this restriction.

To assess the impact of the outcome definition, we repeated all analyses defining depression solely as an EPDS score ≥ 13 (i.e., not including antidepressant use).

Finally, we repeated the multivariable regression analyses using observed age at menopause with additional adjustment for history of depression and antidepressant use in the reproductive period.

All analyses were performed in R (version 4.4.2). Multivariable regression models were run in MLwiN (version 3.13) via the R2MLwiN package (version 0.8.9) using the runMLwiN function.

## Results

### Participant Characteristics

Baseline characteristics were broadly similar across the three analytic samples (Table 1).

**Table 1.**
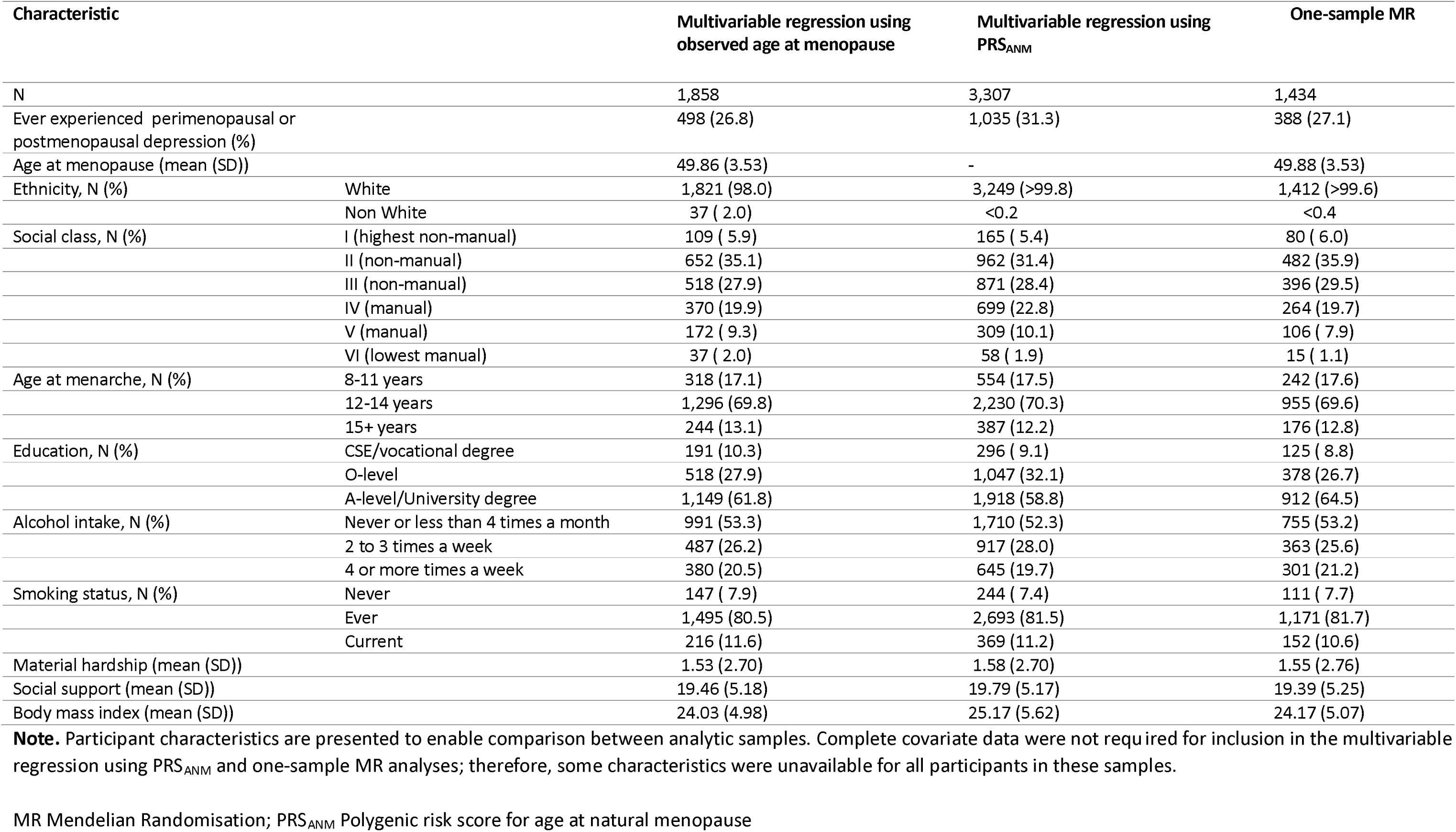
Distribution of covariates at baseline.

| Characteristic |  | Multivariable regression using<br>observed age at menopause | Multivariable regression using<br>PRS<sub>ANM</sub> | One-sample MR |
| --- | --- | --- | --- | --- |
| N |  | 1,858 | 3,307 | 1,434 |
| Ever experienced perimenopausal or<br>postmenopausal depression (%) |  | 498 (26.8) | 1,035 (31.3) | 388 (27.1) |
| Age at menopause (mean (SD)) |  | 49.86 (3.53) | - | 49.88 (3.53) |
| Ethnicity, N (%) | White | 1,821 (98.0) | 3,249 (>99.8) | 1,412 (>99.6) |
|  | Non White | 37 ( 2.0) | <0.2 | <0.4 |
| Social class, N (%) | I (highest non-manual) | 109 ( 5.9) | 165 ( 5.4) | 80 ( 6.0) |
|  | II (non-manual) | 652 (35.1) | 962 (31.4) | 482 (35.9) |
|  | III (non-manual) | 518 (27.9) | 871 (28.4) | 396 (29.5) |
|  | IV (manual) | 370 (19.9) | 699 (22.8) | 264 (19.7) |
|  | V (manual) | 172 ( 9.3) | 309 (10.1) | 106 ( 7.9) |
|  | VI (lowest manual) | 37 ( 2.0) | 58 ( 1.9) | 15 ( 1.1) |
| Age at menarche, N (%) | 8-11 years | 318 (17.1) | 554 (17.5) | 242 (17.6) |
|  | 12-14 years | 1,296 (69.8) | 2,230 (70.3) | 955 (69.6) |
|  | 15+ years | 244 (13.1) | 387 (12.2) | 176 (12.8) |
| Education, N (%) | CSE/vocational degree | 191 (10.3) | 296 ( 9.1) | 125 ( 8.8) |
|  | O-level | 518 (27.9) | 1,047 (32.1) | 378 (26.7) |
|  | A-level/University degree | 1,149 (61.8) | 1,918 (58.8) | 912 (64.5) |
| Alcohol intake, N (%) | Never or less than 4 times a month | 991 (53.3) | 1,710 (52.3) | 755 (53.2) |
|  | 2 to 3 times a week | 487 (26.2) | 917 (28.0) | 363 (25.6) |
|  | 4 or more times a week | 380 (20.5) | 645 (19.7) | 301 (21.2) |
| Smoking status, N (%) | Never | 147 ( 7.9) | 244 ( 7.4) | 111 ( 7.7) |
|  | Ever | 1,495 (80.5) | 2,693 (81.5) | 1,171 (81.7) |
|  | Current | 216 (11.6) | 369 (11.2) | 152 (10.6) |
| Material hardship (mean (SD)) |  | 1.53 (2.70) | 1.58 (2.70) | 1.55 (2.76) |
| Social support (mean (SD)) |  | 19.46 (5.18) | 19.79 (5.17) | 19.39 (5.25) |
| Body mass index (mean (SD)) |  | 24.03 (4.98) | 25.17 (5.62) | 24.17 (5.07) |

**Note.** Participant characteristics are presented to enable comparison between analytic samples. Complete covariate data were not required for inclusion in the multivariable regression using PRS<sub>ANM</sub> and one-sample MR analyses; therefore, some characteristics were unavailable for all participants in these samples.
MR Mendelian Randomisation; PRS<sub>ANM</sub> Polygenic risk score for age at natural menopause

In multivariable regression analyses using observed age at menopause, 1,282 women were included in the perimenopausal analysis and 1,644 women in the postmenopausal analysis, contributing a mean of 1.4 (SD = 0.6) and 1.4 (SD = 0.5) depression assessments, respectively. Overall, 20% and 22% of women ever experienced depression during either the perimenopausal and postmenopausal periods, respectively.

Multivariable regression analyses using PRS_ANM_ as the exposure included 1,997 women in the perimenopausal analysis and 2,767 women in the postmenopausal analysis, contributing a mean of 1.4 (SD = 0.7) and 2.9 (SD = 1.4) depression assessments, respectively. Overall, 20% and 29% of women ever experienced depression during either the perimenopausal and postmenopausal periods, respectively.

One-sample MR analyses included 1,008 women in the perimenopausal analysis and 1,259 women in the postmenopausal analysis, contributing a mean of 1.4 (SD = 0.6) and 1.4 (SD = 0.5) depression assessments, respectively. Overall, 20% and 22% of women ever experienced depression during either the perimenopausal and postmenopausal periods, respectively.

### Multivariable regression using observed age at menopause

There was little evidence that age at menopause was associated with depression during perimenopause (OR per year increase in age at menopause 0.98, 95%CI 0.89 – 1.08) or during postmenopause (OR 1.00, 95%CI 0.93 – 1.06) (Table 2). When perimenopausal and postmenopausal observations were combined (Table 3), there was similarly little evidence of an association (OR 0.99, 95%CI 0.95 – 1.03; N= 1,858).

**Table 2.** Association between age at natural menopause and depression during perimenopausal and postmenopausal periods.

| Analysis type | Exposure |  | Odds ratio | 95% CI | P | N |
| --- | --- | --- | --- | --- | --- | --- |
| **Depression during perimenopause** |  |  |  |  |  |  |
| Multivariable regression | Age at menopause (1-yr increase) | Unadjusted | 1.00 | 0.97, 1.04 | 0.831 | 1,282 |
| Multivariable regression | Age at menopause (1-yr increase) | Adjusted* | 0.98 | 0.89, 1.08 | 0.652 | 1,282 |
| Multivariable regression | PRS<sub>ANM</sub> (1-SD increase) | Adjusted<sup>†</sup> | 0.98 | 0.89, 1.09 | 0.766 | 1,997 |
| One-Sample MR | Age at menopause (1-yr increase) | –<sup>‡</sup> | 1.00 | 0.89, 1.13 | 0.984 | 1,008 |
| **Depression during postmenopause** |  |  |  |  |  |  |
| Multivariable regression | Age at menopause (1-yr increase) | Unadjusted | 0.97 | 0.94, 1.00 | 0.095 | 1,644 |
| Multivariable regression | Age at menopause (1-yr increase) | Adjusted* | 1.00 | 0.93, 1.06 | 0.916 | 1,644 |
| Multivariable regression | PRS<sub>ANM</sub> (1-SD increase) | Adjusted<sup>†</sup> | 0.92 | 0.86, 0.99 | 0.036 | 2,767 |
| One-Sample MR | Age at menopause (1-yr increase) | –<sup>‡</sup> | 0.97 | 0.86, 1.09 | 0.568 | 1,259 |

\*Adjusted for age, ethnicity, social class, education, age at menarche, material hardship, social support, smoking status, body mass index and alcohol intake.
<sup>†</sup>Adjusted for age and the first 10 principal components of ancestry
<sup>‡</sup>Adjusted for the first 10 principal components of ancestry
CI: confidence interval; MR: Mendelian randomization, PRS<sub>ANM</sub>: Polygenic risk score for age at natural menopause, SD: standard deviation

**Table 3.**
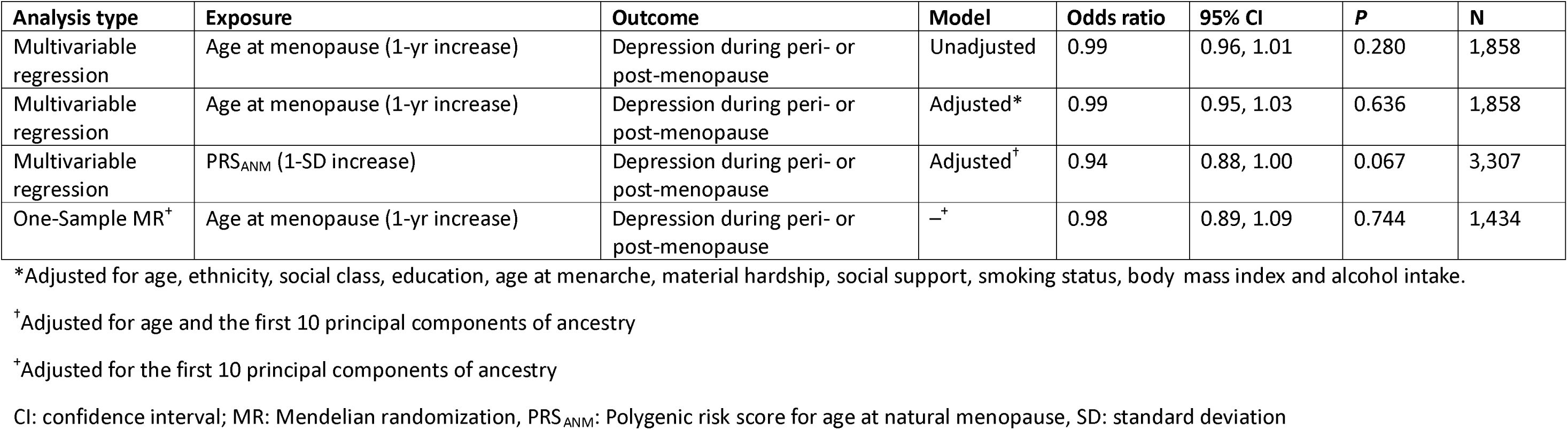
Association between age at natural menopause and depression during peri- or post-menopause.

| Analysis type | Exposure | Outcome | Model | Odds ratio | 95% CI | P | N |
| --- | --- | --- | --- | --- | --- | --- | --- |
| Multivariable regression | Age at menopause (1-yr increase) | Depression during peri- or post-menopause | Unadjusted | 0.99 | 0.96, 1.01 | 0.280 | 1,858 |
| Multivariable regression | Age at menopause (1-yr increase) | Depression during peri- or post-menopause | Adjusted* | 0.99 | 0.95, 1.03 | 0.636 | 1,858 |
| Multivariable regression | PRS<sub>ANM</sub> (1-SD increase) | Depression during peri- or post-menopause | Adjusted<sup>†</sup> | 0.94 | 0.88, 1.00 | 0.067 | 3,307 |
| One-Sample MR<sup>‡</sup> | Age at menopause (1-yr increase) | Depression during peri- or post-menopause | —<sup>‡</sup> | 0.98 | 0.89, 1.09 | 0.744 | 1,434 |

\*Adjusted for age, ethnicity, social class, education, age at menarche, material hardship, social support, smoking status, body mass index and alcohol intake.
<sup>†</sup>Adjusted for age and the first 10 principal components of ancestry
<sup>‡</sup>Adjusted for the first 10 principal components of ancestry
CI: confidence interval; MR: Mendelian randomization, PRS<sub>ANM</sub>: Polygenic risk score for age at natural menopause, SD: standard deviation

### Multivariable regression using genetic proxy for age at menopause (PRS_ANM_)

Among the PRSs constructed using multiple GWAS *p*-value thresholds, a threshold of *p* ≤ 0.1 was selected for the main analysis as it explained the greatest variance in age at menopause (R^2^ = 13%, eTable 4, eFigure 1), while balancing predictive power against the increased risk of potential pleiotropy at more liberal thresholds. This PRS comprised 47,699 independent SNPs. In ALSPAC women with estimated age at menopause available, the PRS was positively associated with age at menopause (β = 1.3 years per SD increase in PRS; SE = 0.08; *p* = 2.1x10^-51^).

There was little evidence of an association between PRS_ANM_ and depression during perimenopause (OR per SD increase in PRS_ANM_ 0.98, 95%CI 0.89 – 1.09) (Table 2). However, there was evidence of an association during postmenopause, with higher genetically predicted age at menopause associated with lower risk of depression (OR 0.92, 95%CI 0.86 – 0.99). When menopausal stages were combined, there was weak evidence of an association (OR 0.94, 95%CI 0.88 – 1.00; N = 3,307)(Table 3). Findings were consistent across different PRS p-value thresholds (eTable 5).

#### One-sample Mendelian Randomisation

The PRS explained 8% of the variance in age at menopause (F-statistic = 165), indicating a sufficiently strong instrument. In this analysis, there was little evidence that age at menopause had a causal effect on depression during perimenopause (OR per year increase in age at menopause 1.00, 95%CI 0.89 – 1.13) or postmenopause (OR 0.97, 95%CI 0.86 – 1.09) (Table 2). When stages were combined, there was similarly little evidence of a causal effect (OR 0.98, 95%CI 0.89 – 1.09; N = 1,434) (Table 3).

#### Sensitivity analyses

Results were consistent when including all postmenopausal observations (i.e., not restricted to within 8 years of menopause) (eTable 6). In these analyses, depression assessments extended up to 30 years after menopause.

Results remained similar after additional adjustment for history of depression and antidepressant use during the reproductive period (eTable 7) and were consistent when depression was defined using only EPDS ≥ 13 (eTable 8).

## Discussion

In this prospective cohort study, effect estimates across multivariable regression and genetic analyses were generally directionally consistent with a small inverse association between age at menopause and risk of depression during the perimenopausal and early postmenopausal periods. However, associations were small and most estimates spanned the null, suggesting that any effect is likely modest and that menopausal timing is unlikely to be a major determinant of midlife depression risk. Despite differing assumptions and sources of bias across analytic methods, the broadly consistent direction of effect across analytic approaches provides some support for a small association between later menopause and lower depression risk. More broadly, the consistency observed across complementary analytic approaches highlights the value of triangulating evidence, particularly where individual methods alone are inconclusive.

Key strengths include the large community-based prospective cohort, repeated measures of depressive symptoms and antidepressant use across the menopausal transition, and the use of complementary analytic approaches, including genetic methods, to reduce confounding and reverse causation.

Several limitations should also be considered. Analyses were restricted to women with an estimated age at menopause or known menopausal stage. Women included in the analyses were more socioeconomically advantaged and healthier than those excluded due to missing information on age at menopause or menopausal stage (eTable 9), which may limit generalisability and introduce selection bias. Power may have been limited for detecting small effects particularly during perimenopause where depressive symptom assessments were less frequent and sample sizes were smaller. Missing data is another consideration. Although missingness in confounders was generally low (eTable 10), with the highest proportion observed for social class (7.7%), we did not impute missing confounder data due to the complexity of the multilevel longitudinal structure. Instead, we used a complete-case approach, whereby analyses included only participants with complete data on variables included in the model. This resulted in only a modest reduction in sample size, but bias due to data not missing at random cannot be excluded. Finally, the cohort predominantly comprised White European, parous women, limiting generalisability to more ethnically diverse and nulliparous populations.

Our findings are broadly consistent with previous observational studies. A meta-analysis of 67,714 women^14^ reported that each two-year increase in age at menopause was associated with a 2% lower odds of postmenopausal depression (OR 0.98; 95%CI 0.96-0.99). Although our analyses using observed age at menopause provided little evidence of an association during postmenopause, analyses using a PRS for age at menopause as an exposure suggested lower postmenopausal depression risk, overall providing some support for a small inverse association in postmenopause.

Notably, the effect sizes reported in the meta-analysis were small and much of the contributing evidence was cross-sectional.

The Study of Women’s Health Across the Nation (SWAN)^15^, a longitudinal, multiethnic, multisite, community-based study of menopause in the US found that older age at onset of the perimenopause was associated with a 6% lower odds of depression during the transition and up to 10 years postmenopause (OR 0.94 per year increase in age at menopause, 95%CI 0.90-0.97, N=5,695). In our study, when perimenopausal and postmenopausal observations were analysed jointly, effect estimates were directionally consistent with a modest protective association, aligning with these findings despite differences in exposure definition.

Overall, our findings support that menopausal timing itself is unlikely to be a major determinant of midlife depression risk. However, the consistent direction of effects across analyses suggest that a small inverse association may exist. Vulnerability to depression during midlife may instead relate more strongly to hormonal fluctuations, menopausal symptoms (such as vasomotor symptoms^39^ and sleep disturbances^40^), and broader psychosocial stressors and physical health changes during this life stage. Later menopause is socially patterned and associated with better overall health, higher socioeconomic position, and healthier behaviours (e.g. lower rates of smoking)^41^, factors that may confound observational associations and influence how the menopausal transition is experienced.

## Conclusion

In this large, prospective cohort study combining conventional multivariable regression and genetic approaches, we found limited evidence that age at menopause is associated with depression during the perimenopausal and early postmenopausal years. Effect estimates were small but directionally consistent across the analytic methods, suggesting that any causal effect, if present, is likely small. Taken together these findings suggest that menopausal timing alone is unlikely to be a major driver of midlife depression risk.

Future studies with larger samples, more precise characterisation of menopausal stage, and repeated measures of both hormonal and mental health outcomes will be important to clarify the extent and timing of any causal effects. Improved understanding may inform more targeted monitoring and prevention strategies around the menopausal transition.

## Data Availability

The informed consent obtained from ALSPAC (Avon Longitudinal Study of Parents and Children) participants does not allow the data to be made available through any third party maintained public repository. Supporting data are available from ALSPAC on request under the approved proposal number, B4425. Full instructions for applying for data access can be found here: http://www.bristol.ac.uk/alspac/researchers/access/. The ALSPAC study website contains details of all available data (http://www.bristol.ac.uk/alspac/researchers/our-data/).
Any questions regarding data or sample access should be directed to (data) or (samples).
All code used to run analyses are available on GitHub: RochelleKnight/Association-of-age-at-menopause-and-risk-of-depression-during-the-perimenopause-and-postmenopause
Archived software available from: https://doi.org/10.5281/zenodo.21358939

https://github.com/RochelleKnight/Association-of-age-at-menopause-and-risk-of-depression-during-the-perimenopause-and-postmenopause

## Declarations

### Ethical approval and consent

Ethical approval for the study was obtained from the ALSPAC Ethics and Law Committee and the Local Research Ethics Committees. Informed consent for the use of all data collected was obtained from participants following the recommendations of the ALSPAC Ethics and Law Committee at the time. Study participation is voluntary and during all data collection sweeps, information was provided on the intended use of data. Participants can contact the study team at any time to retrospectively withdraw consent for their data to be used. The completion of a questionnaire, either on paper or online, was considered to be written consent from participants to use their data for research purposes. Full details of the approvals are available from the study website.

### Availability of Data and Materials

The informed consent obtained from ALSPAC (Avon Longitudinal Study of Parents and Children) participants does not allow the data to be made available through any third party maintained public repository. Supporting data are available from ALSPAC on request under the approved proposal number, B4425. Full instructions for applying for data access can be found here: http://www.bristol.ac.uk/alspac/researchers/access/. The ALSPAC study website contains details of all available data (http://www.bristol.ac.uk/alspac/researchers/our-data/).

Any questions regarding data or sample access should be directed to (data) or (samples).

All code used to run analyses are available on GitHub: RochelleKnight/Association-of-age-at-menopause-and-risk-of-depression-during-the-perimenopause-and-postmenopause

Archived software available from: https://doi.org/10.5281/zenodo.21358939

## Author Contributions

**Rochelle Knight:** Conceptualization, Methodology, Software, Data Curation, Writing – Original Draft. **Carol Joinson:** Conceptualization, Methodology, Writing - Review & Editing, Supervision. **Abigail Fraser:** Conceptualization, Methodology, Writing - Review & Editing, Supervision. **Kimberly Burrows:** Methodology, Software, Data Curation, Writing - Review & Editing. **Ana Gonçalves Soares:** Conceptualization, Methodology, Software, Writing - Review & Editing, Supervision

## Acknowledgements

We are extremely grateful to all the families who took part in this study, the midwives for their help in recruiting them, and the whole ALSPAC team, which includes data collection staff, data and administrations staff, technical managers and the technical staff with the Bristol Bioresource Laboratory, based within the University of Bristol.

