## Supplementary Methods for "Association of age at menopause and risk of depression during the perimenopause and postmenopause"

**Estimation of age at natural menopause and reproductive stage**

Age at natural menopause was estimated using self-reported menstrual bleeding data, as described in detail elsewhere^1^. Briefly, women provided information on their menstrual cycles at up to eight questionnaire timepoints. At each timepoint they reported:

i. The date of their last menstrual period (LMP) (can be reported incompletely);
ii. Whether they had experienced a menstrual period in the previous 3 months (yes/no);
iii. Whether they had experienced a menstrual period in the previous 12 months (yes/no).

Where possible, the date of the LMP was estimated for each timepoint. The final menstrual period (FMP) was defined as the last reported menstrual period followed by at least 12 consecutive months of amenorrhea. Age at menopause was defined as the age at the FMP.

**Assignment of reproductive stage**

At each of the eight timepoints, women were classified into one of three mutually exclusive reproductive stages based on menstrual bleeding patterns, in accordance with the STRAW^2^ criteria:

i. Reproductive (STRAW category -4, -3b and -3a)
ii. Perimenopause (STRAW category -2, -1 and +1a)
iii. Postmenopause (STRAW category ≥ +1b, irrespective of the years since menopause))

This approach allowed inclusion of women without an estimated ANM. For example, women who could not recall the date of their last menstrual period but reported no bleeding in the preceding 12 months could be classified as postmenopausal. Similarly, women who had not yet reached menopause by the final follow-up could still be classified as perimenopausal based on bleeding patterns.

STRAW staging was applied at each timepoint, with consistency checks to ensure biologically plausible progression through reproductive stages. Because reproductive stage was determined using menstrual bleeding patterns, factors known to influence menstruation were considered, including surgery to reproductive organs, use of hormonal contraception or hormone replacement therapy (HRT), and other causes such as chemotherapy, radiotherapy, endometrial ablation, pregnancy, or breastfeeding. Full details of censoring and exclusions are reported elsewhere^1^.

**Definition of recent menstrual activity**

To distinguish the reproductive from the perimenopausal period, menstrual activity was classified as:

- **Recent period:** LMP within the last 60 days
- **No recent period:** LMP is ≥ 60 days and ≤ 365 days ago

Information on cycle regularity was also collected. Please see below.

| ALSPAC Variable | Question(s)/measure(s) | Responses | Final variable coding |
| --- | --- | --- | --- |
| Period regularity |  |  |  |
| fm1ob130, fm2ob130, fm3ob130, fm4ob130 | Are your periods regular? | 1: Yes, every 28-30 days  2: Yes, < every 28 days  3: Yes, every 30 days  4: No | Recode (4=0) (1-3 = 1)  0: Irregular periods  1: Regular periods |
| t4837, V4837 | Please describe your most recent periods:  Are your periods irregular? | 1: Very  2: Moderately  3: Mildly  4: Not at all | Recode (1/2 = 0) (3/4=0)  0: Irregular periods  1: Regular periods |
| U1050 | Are your periods regular? | 1: Yes, every 23 days or less  2: Yes, every 24-35 days  3: Yes, > every 35 days  4: No | Recode (1-3 = 1) (4=0)  0: Irregular periods  1: Regular periods |

**Assigning the reproductive and perimenopausal period**

Menstrual activity and cycle regularity were jointly used to distinguish the reproductive from the perimenopausal period at each timepoint as shown below.

| Period regularity | Recent period | No recent period |
| --- | --- | --- |
| Regular | Reproductive | Perimenopause |
| NA | NA | Perimenopause |
| Irregular | Perimenopause | Perimenopause |

Where ANM was available, perimenopause was additionally defined as the period spanning from three years prior to ANM up to one year after ANM. Within the three years prior to ANM, any timepoints previously assigned as the reproductive period were retained.

**Assigning the postmenopausal period**

Where ANM was available, timepoints that were >1 year post-ANM were assigned as postmenopausal. Where ANM was not available, women reporting no menstrual period in the preceding 12 months (yes/no response options) were classified as postmenopausal at that timepoint.

**Additional data on menopausal status**

In one questionnaire (Questionnaire MB), reproductive stage status was assessed using a different set of questions. Women were asked:

- Which of the following statements best describes your current
- menopause status?:
  1. I have been through the menopause
  2. I am going through the menopause now
  3. I have not yet started going through the menopause
  4. I am not sure
- When their last menstrual period had occurred:
  1. Within the past 3 months
  2. 4-12 months ago
  3. More than 12 months ago

Women were not assigned a reproductive stage if they reported:

- Their period had stopped as they had a hysterectomy
- Their period had stopped for other or unknown reasons
- They were pregnant or breastfeeding

|  | Period in last 3 months | Period 4-12 months ago | Period more than 12 months ago |
| --- | --- | --- | --- |
| Not started going through the menopause | Reproductive | Perimenopause | Perimenopause |
| Going through the menopause | Perimenopause | Perimenopause | Postmenopause |
| Been through the menopause | NA | NA | Postmenopause |

Reproductive stage for this questionnaire was then assigned using the classification shown below.

Women reporting the use of current contraceptives or HRT were assigned a reproductive stage only based on their response to their current menopausal status.

**Consistency checks**

The above process resulted in women having up to nine timepoints with an assigned reproductive stage. These timepoints were then checked to ensure that each woman’s reproductive status progressed in the correct chronological order over time.

**Consistency check 1:** Woman is classified as reproductive after being classified as perimenopausal

Scenario 1.1

A woman is classified as being in the perimenopause, then classified as reproductive at the next timepoint. If, for at least the two subsequent timepoints, she is classified as either in the perimenopause or postmenopause, her earlier classification of reproductive is changed to perimenopause.

| Timepoint 1 | Timepoint 2 | Timepoint 3 | Timepoint 4 | Timepoint 5 |
| --- | --- | --- | --- | --- |
| Perimenopause | Reproductive | Perimenopause | Perimenopause | Postmenopause |
| Perimenopause | Reproductive | Postmenopause | Postmenopause | Postmenopause |
| Reproductive | Perimenopause | Reproductive | Perimenopause | Postmenopause |

Scenario 1.2

A woman is classified as being in the perimenopause for two subsequent timepoints, then classified as reproductive. The timepoint following the reproductive classification is either perimenopause, postmenopause or there are no further timepoints with STRAW classification. Her classification of reproductive is changed to perimenopause.

| Timepoint 1 | Timepoint 2 | Timepoint 3 | Timepoint 4 | Timepoint 5 |
| --- | --- | --- | --- | --- |
| Perimenopause | Perimenopause | Reproductive | NA |  |
| Perimenopause | Perimenopause | Reproductive | Perimenopause | NA |
| Perimenopause | Perimenopause | Reproductive | Postmenopause | NA |

Scenario 1.3

A women is classified as being in the perimenopause, then for the following two timepoints as reproductive. The classification of perimenopause is changed to reproductive.

| Timepoint 1 | Timepoint 2 | Timepoint 3 | Timepoint 4 | Timepoint 5 |
| --- | --- | --- | --- | --- |
| Perimenopause | Reproductive | Reproductive | Perimenopause | Perimenopause |
| Perimenopause | Reproductive | Reproductive | Reproductive | Perimenopause |
| Reproductive | Perimenopause | Reproductive | Reproductive | NA |

Scenario 1.4

A women is classified as being in the perimenopause, then classified as reproductive at the next timepoint. There are no further timepoints classified as reproductive. All timepoints prior to the perimenopause classification are either reproductive or the perimenopause classification was the first timepoint available with a STRAW classification. The final reproductive classification is either only followed by one further timepoint with a classification of perimenopause or postmenopause, or is the final timepoint with a STRAW classification.

| Timepoint 1 | Timepoint 2 | Timepoint 3 | Timepoint 4 | Timepoint 5 |
| --- | --- | --- | --- | --- |
| Reproductive | Perimenopause | Reproductive | NA |  |
| Reproductive | Perimenopause | Reproductive | Perimenopause | NA |
| Reproductive | Perimenopause | Reproductive | Postmenopause | NA |
| Perimenopause | Reproductive | NA |  |  |
| Perimenopause | Reproductive | Perimenopause | NA |  |
| Perimenopause | Reproductive | Postmenopause | NA |  |

If the women’s age at the reproductive timepoint is ≤ 3 years below her age at menopause or the population mean age of menopause if ANM is unavailable, the classification of reproductive should be changed to perimenopause. If the reproductive timepoint is followed a postmenopause classification or is the final available timepoint with a straw classification, and the women’s age at the reproductive timepoint is greater than the population mean age of menopause, the classification of reproductive should be changed to postmenopause. Else the perimenopause timepoint should be changed to reproductive.

**Consistency check 2**: Woman is classified as reproductive or perimenopausal after being classified as postmenopause.

Scenario 2.1

A women is classified as being postmenopause, then as either reproductive or perimenopausal at the following timepoint. There are no further timepoints with a reproductive stage classification. This will **only** occur when there is no ANM available for the women and we only have information that she has not bled in the last 12 months. The woman then comes back at a later timepoint and says she has had another bleed.

If the women’s age at the reproductive timepoint is less than the population mean age of menopause, the postmenopause classification is changed to reproductive. If the women’s age at the reproductive timepoint is greater than the population mean age of menopause, the reproductive timepoint is changed to postmenopause.

If the women’s age at the perimenopause timepoint is less than the population mean age of menopause, the postmenopause classification is changed to perimenopause. If the women’s age at the perimenopause timepoint is greater than the population mean age of menopause, the reproductive timepoint is changed to postmenopause.

| Timepoint 1 | Timepoint 2 | Timepoint 3 | Age at Timepoint 3 | Change to |
| --- | --- | --- | --- | --- |
| Postmenopause | Reproductive | NA | 45 | Postmenopause to reproductive |
| Postmenopause | Reproductive | NA | 51 | Reproductive to postmenopause |
| Postmenopause | Perimenopause | NA | 45 | Postmenopause to perimenopause |
| Postmenopause | Perimenopause | NA | 51 | Perimenopause to postmenopause |

**Consistency check 3**: Woman is classified as postmenopause prior to being classified as perimenopause or reproductive.

Scenario 3.1

| Timepoint 1 | Timepoint 2 | Timepoint 3 | Change to |
| --- | --- | --- | --- |
| Postmenopause | Reproductive | Reproductive | Reproductive |
| Postmenopause | Perimenopause | Perimenopause | Perimenopause |

A woman is classified as being postmenopause, then classified as reproductive or perimenopause for the following two timepoints. Her classification of postmenopause should be changed to reproductive or perimenopause.

**Consistency check 4**: Woman switches between reproductive and perimenopause.

Scenario 4.1

For all further scenarios in which a woman switches between perimenopause and reproductive and we have been unable to change inaccuracies based on any of the above, we assign reproductive stage based on age and time to menopause.

If the women’s age at the timepoint is more than 3 years below her ANM or population mean age of menopause, she is classified as reproductive. Otherwise she is classified as perimenopause.

Example:

| Timepoint 1 | Timepoint 2 | Timepoint 3 | Timepoint 4 | Timepoint 5 | Timepoint 6 |
| --- | --- | --- | --- | --- | --- |
| Reproductive | Perimenopause | Reproductive | Perimenopause | Reproductive | Menopause transition |

The columns highlighted in yellow are assigned menopausal status based on age at timepoint. Timepoints 1 and 6 are not changed.

**Assigning reproductive stage to EPDS assessments**

Reproductive stage was assigned at up to nine timepoints based on menstrual bleeding patterns and classified according to STRAW criteria, as described above. The EPDS was measured at 11 separate questionnaires, which did not always coincide with when the menstrual questions were asked. Therefore, reproductive stage for each EPDS assessment was assigned using information from the nearest observed reproductive stage classification.

For each women, the observed reproductive stage classifications were first used to identify:

- The latest timepoint classified as reproductive
- The earliest and latest timepoints classified as perimenopausal
- The earliest timepoint classified as postmenopausal

Where a women had only a single observed perimenopausal timepoint, a ±6-month window around that assessment was used to assign EPDS timepoints.

Using these boundaries, reproductive stage was assigned to each EPDS timepoint as follows:

- **Reproductive**: EPDS assessments occurring before the latest observed reproductive stage timepoint.
- **Perimenopausal**: EPDS assessments occurring between the first and last observed perimenopausal timepoints (or within ±6-months where only one perimenopausal observation was available).
- **Postmenopausal**: EPDS assessments occurring after the earliest observed postmenopausal timepoint.

Additionally, where ANM was available, perimenopause was defined as the period spanning from three years prior to ANM up to one year after ANM, and postmenopausal thereafter (>1 year post-ANM). Within the three years prior to ANM, any previously assigned reproductive status was retained.

**Confounders**

Ethnicity, social class, age at menarche, and educational attainment were treated as baseline confounders. These were collected around the time of recruitment (mean age 28.6 years, SD 4.9). Missing responses for ethnicity, age at menarche, and education were obtained where possible from subsequent questionnaires. Social class and history of depression were only recorded at baseline. Material hardship, social support, smoking status, BMI, and alcohol intake were treated as time-varying confounders. For each depression assessment, the most recent non-missing response prior to or at that time point was used.

**Genotyping of ALSPAC Mothers**

The ALSPAC mothers and their children were genotyped as one dataset. We include how the ALSPAC children were genotyped for completeness however only the ALSPAC mothers were used in this analysis.

Associated publications:

ALSPAC children: Horikoshi et al 2013^3^

ALSPAC mothers: Rietveld et al 2013^4^

ALSPAC children were genotyped using the Illumina HumanHap550 quad chip genotyping platform. The resulting raw genome-wide data were subjected to standard quality control methods. Individuals were excluded on the basis of gender mismatches; minimal or excessive heterozygosity; disproportionate levels of individual missingness (>3%) and insufficient sample replication (IBD < 0.8). Population stratification was assessed by multidimensional scaling analysis and compared with Hapmap II (release 22) European descent (CEU), Han Chinese, Japanese and Yoruba reference populations; all individuals with non-European ancestry were removed. SNPs with a minor allele frequency of < 1%, a call rate of < 95% or evidence for violations of Hardy-Weinberg equilibrium (P < 5E-7) were removed. Cryptic relatedness was measured as proportion of identity by descent (IBD > 0.1). This resulted in 9,115 subjects and 500,527 SNPs passing QC.

Genetic data for ALSPAC mothers were generated using the Illumina human660W-Quad Array. Quality control was carried out on an initial sample of 10,015 women and 557,124 directly genotyped single nucleotide polymorphisms (SNPs) using PLINK v1.07. SNPs were removed if they displayed more than 5% missingness or a Hardy-Weinberg equilibrium P value of less than 1.0e-06. Additionally, SNPs with a minor allele frequency of less than 1% were removed. Samples were excluded if they displayed more than 5% missingness, had indeterminate X chromosome heterozygosity or extreme autosomal heterozygosity. Samples showing evidence of population stratification were identified by multidimensional scaling of genome-wide identity by state pairwise distances using the four HapMap populations as a reference and then excluded.

Cryptic relatedness was assessed using a IBD estimate of more than 0.125 which is expected to correspond to roughly 12.5% alleles shared IBD or a relatedness at the first cousin level. Related subjects that passed all other quality control thresholds were retained. This resulted in 9,048 subjects and 526,688 SNPs passed these quality control filters.

There were 477,482 SNP genotypes in common between the sample of mothers and sample of children that were combined. SNPs with genotype missingness above 1% due to poor quality were removed (11,396 SNPs removed) and a further 321 subjects were removed due to potential ID mismatches. This resulted in a dataset of 17,842 subjects containing 6,305 duos and 465,740 SNPs (112 were removed during liftover and 234 were out of HWE after combination).

Haplotypes were estimated using ShapeIT (v2.r644) which utilises relatedness during phasing.

Imputation of the target data was performed using Impute V2.2.2 against the 1000 genomes reference panel (Phase 1, Version 3). This gave 8,196 eligible mothers with available genotype data after exclusion of related subjects using cryptic relatedness measures described previously.

After removing those who had withdrawn consent, the final sample comprised 7,728 unrelated mothers with genome-wide genetic data, including approximately 8.1 million SNPs after imputation.

**Derivation of PRS for age at menopause**

Prior to PRS derivation, ambiguous SNPs (i.e., complementary alleles C/G or A/T) and duplicate SNPs were removed from the GWAS summary statistics, resulting in a cleaned dataset of 11,290,199 SNPs. SNPs were mapped to the same genome build used in ALSPAC (GRCh37/hg19), of which 6,069,420 were present in both the GWAS summary statistics and the ALSPAC mothers’ genotype data.

PRS_ANM_ were generated across multiple GWAS p-value thresholds using PRSice-2. Linear regression models were fitted for each threshold, adjusting for the first 10 principal components (PCs) of ancestry to assess the association between PRS_ANM_ and age at natural menopause (ANM). Model performance was assessed using R^2^, calculated as the difference between the R^2^ of the full model (ANM ~ PRS + PC1–PC10) and the null model (ANM ~ PC1–PC10) (Supplementarty Figure 1, Supplementary Table 4).

Although PRS performance generally increased at more liberal GWAS p-value thresholds due to the inclusion of additional SNPs, these thresholds may also increase the likelihood of incorporating SNPs unrelated to ANM through horizontal pleiotropy. Therefore, selection of the best-fitting PRS_ANM_ used in the subsequent analyses was informed by comparison of model performance across thresholds via R^2^ (Supplementary Figure 1, Supplementary Table 4) alongside consideration for potential pleiotropy.

1.	Knight R, Fraser A, Joinson C, Soares A. Estimating age of menopause in mothers in the ALSPAC Study: A data note [version 1; peer review: awaiting peer review]. *Wellcome Open Research*. 2025;10(631)doi:10.12688/wellcomeopenres.24760.1

2.	Harlow SD, Gass M, Hall JE, et al. Executive summary of the Stages of Reproductive Aging Workshop + 10. *Menopause*. 2012;19(4):387-395. doi:10.1097/gme.0b013e31824d8f40

3.	Horikoshi M, Yaghootkar H, Mook-Kanamori DO, et al. New loci associated with birth weight identify genetic links between intrauterine growth and adult height and metabolism. *Nature Genetics*. 2013/01/01 2013;45(1):76-82. doi:10.1038/ng.2477

4.	Rietveld CA, Medland SE, Derringer J, et al. GWAS of 126,559 Individuals Identifies Genetic Variants Associated with Educational Attainment. *Science*. 2013;340(6139):1467-1471. doi:doi:10.1126/science.1235488
