## Supplementary Tables and Figures for "Association of age at menopause and risk of depression during the perimenopause and postmenopause"

**Association of age at menopause and risk of depressive symptoms during the menopause transition and postmenopause**

**Supplementary Tables and Figures**

**Supplementary Tables**

**Supplementary Figures**

eTable 1. Derivation of the Edinburgh Post Natal Depression Scale (EPDS) depressive symptom score outcome variable from Avon Longitudinal Study of Parents and Children (ALSPAC).

| **Data collection** | **ALSPAC Variable** | **Question(s)/measure(s)** | **Responses** | **Final variable coding** |
| --- | --- | --- | --- | --- |
| **EPDS** | | | | |
|  |  | **In the past week** | | Recode  (a), (b), (d): (1 = 0) (2 = 1) (3 = 2) (4 = 3)  (c),(e),(f),(g),(h),(i),(j): (1 = 3) (2 = 2) (3 = 1) (4 = 0)  Summed scored then calculated  **Mode imputation:** Any missing items were put to the mode for that item. If all items were missing, score was set as NA.  (Higher score = higher depressive symptom burden) |
| **Edinburgh Post Natal Depression Scale (EPDS) developed by Cox et al (1987).** | g280, h190, k3030, l2010, n6060, r4010, t3240, V5158, Y4000, covid5m_4000, MB6010, | (a) I have been able to laugh and see the funny side of things | 1: As much as I always could  2: Not quite so much now  3: Definitely not so much now  4:Not at all |  |
|  | g281, h191, k3031, l2011, n6061, r4011, t3241, V5159, Y4010, covid5m_4001, MB6020 | (b) I have looked forward with enjoyment to things | 1: As much as I ever did  2: Rather less than I used to  3: Definitely less than I used to  4: Hardly at all |  |
|  | g282, h192, k3032, l2012, n6062, r4012, t3242, V5150, Y4020, covid5m_4002, MB6030 | (c) I have blamed myself unnecessarily when things went wrong | 1: Yes, most of the time  2: Yes, some of the time  3: Not very often  4: No never |  |
|  | g283, h193, k3033, l2013, n6063, r4013, t3243, V5151, Y4030, covid5m_4003, MB6040 | (d) I have been anxious or worried for no good reason | 1: No, not at all  2: Hardly ever  3: Yes, sometimes  4: Yes, often |  |
|  | g284, h194, k3034, l2014, n6064, r4014, t3244, V5152, Y4040, covid5m_4004, MB6050 | (e) I have felt scared or panicky for no very good reason | 1: Yes, quite a lot  2: Yes, sometimes  3: No, not much  4: No, not at all |  |
|  | g285, h195, k3035, l2015, n6065, r4015, t3245, V5153, Y4050, covid5m_4005, MB6060 | (f) Things have been getting on top of me | 1: Yes, most of the time I haven't been able to cope  2: Yes, sometimes I haven't been coping as well as usual  3: No, most of the time I have coped quite well  4: No, I have been coping as well as ever |  |
|  | g286, h196, k3036, l2016, n6066, r4016, t3246, V5154, Y4060, covid5m_4006, MB6070 | (g) I have been so unhappy that I have had difficulty sleeping | 1: Yes, most of the time  2: Yes, sometimes  3: Not very often  4: No, not at all |  |
|  | g287, h197, k3037, l2017, n6067, r4017, t3247, V5155, Y4070, covid5m_4007, MB6080 | (h) I have felt sad or miserable | 1: Yes, most of the time  2: Yes, quite often  3: Not very often  4: No, not at all |  |
|  | g288, h198, k3038, l2018, n6068, r4018, t3248, V5156, Y4080, covid5m_4008, MB6090 | (i) I have been so unhappy that I have been crying | 1: Yes, most of the time  2: Yes, quite often  3: Only occasionally  4: No, never |  |
|  | g289, h199, k3039, l2019, n6069, r4019, t3249, V5157, Y4090, covid5m_4009, MB6100 | (j) The thought of harming myself has occurred to me | 1: Yes, quite often  2: Sometimes  3: Hardly ever  4: Never |  |

eTable 2. Derivation of current antidepressant use outcome variable from Avon Longitudinal Study of Parents and Children (ALSPAC).

| **Data collection timepoint** | **ALSPAC Variable** | **Question** | **Response** | **Final variable coding** |
| --- | --- | --- | --- | --- |
| G  (The questionnaire was sent to the mother 21 months after the child was born) | g049 | Since your toddler was 8 months old how often have you taken the following?  Option: pills for depression | 1: Every day  2: Often  3: Sometimes  4: Not at all | Recode  (1 = 1) and (2 = 1)  (3 = 0) and (4 = 0)  1: Current antidepressant use  0: No current antidepressant use  Note: Questionnaire S did not have the option to respond “Not at all”. Therefore no response to the question was assumed to correspond to no current antidepressant use. |
| H  (The questionnaire was sent to the mother 33 months after the child was born) | h039 | Since your study child was 18 months old how often have you taken the following?  Option: pills for depression | 1: Every day  2: Often  3: Sometimes  4: Not at all |  |
| J | j044 | In the past year how often have you taken or used the following?  Option: pills for depression | 1: Every day  2: Often  3: Sometimes  4: Not at all |  |
| K | k1044 | In the past year how often have you taken the following?  Option: pills for depression | 1: Every day  2: Often  3: Sometimes  4: Not at all |  |
| L  (The questionnaire was sent to the mother 6 years 1 month after the child was born) | l3044 | Since your study child’s 5th birthday how often have you taken the following?  Option: pills for depression | 1: Every day  2: Often  3: Sometimes  4: Not at all |  |
| P | p1054 | In the last 2 years how often have you taken the following?  Option: pills for depression | 1: Every day  2: Often  3: Sometimes  4: Not at all |  |
| S | s4103 | In the past year how often have you taken or used the following?  Option: pills for depression | 1: Every day  2: Most days  3: Sometimes  4: Once or Twice |  |

eTable 3. Derivation of the confounder variables from Avon Longitudinal Study of Parents and Children (ALSPAC).

| **Data collection** | **ALSPAC Variable** | **Question(s)/measure(s)** | **Responses** | **Final variable coding** |
| --- | --- | --- | --- | --- |
| **Age** | | | | |
|  | g994, h992, k9996a, l9996a, n9992, r9996a, t9994, V9996, Y9992, covid5m_9650, MB9510 | Derived from questionnaire date of completion and participants date of birth | Day/Month/Year | Age (years) |
| **Social Class (antenatal questionnaire)** | | | | |
| 1991 British Office of Population and Census Statistics job codes | C755 | Derived variable from questions:   - Actual job, occupation, trade or profession - Please tick which of the following apply to you: foreman, manager, supervisor, leading hand, self-employed, none of these   Type of industry or service given (main things done in job) | 1: I  2: II  3: III (non-manual)  4: IV (manual)  5: V  6: VI  65: Armed forces  -1: missing | Select highest social class between participant and their partner. Armed forces coded as missing. Coded from 1 (highest) to 6 (lowest).  Definitions:  Non-manual: professional, managerial, or skilled professions  Manual: partly or unskilled occupations |
|  | C765 |  |  |  |
| **Ethnicity (antenatal questionnaire)** | | | | |
|  | c800, n4180 | How would you describe the race or ethnic group of yourself? | -1 Missing  1 White  2 Black Caribbean  3 Black African  4 Other black  5 Indian  6 Pakistani  7 Bangladeshi  8 Chinese  9 Other  99 DK | Recode:  0: White  1: Non-white  Missing values from Questionnaire C were replaced using responses from Questionnaire N |
| **Education** | | | | |
|  | c645a | Derived variable: mother’s highest educational qualification | 1: CSE/none  2: Vocational  3: O-level  4: A-level  5: Degree  -1: Missing | Recode:  0: A-level or greater  1: O-level  2: Vocational or less  Education variables were not derived by ALSPAC for Questionnaires K and N. The same coding as applied in questionnaire C was used.  The highest educational qualification reported was used. It was felt that a lot of mothers with no educational qualifications left this whole question blank. Therefore, all women who did not respond to any of the questions were coded to 2 (Vocational or less) |
|  | k6280, k6281 | CSE/none |  |  |
|  | k6284, k6285, k6288, k6295 | Vocational |  |  |
|  | k6282, | O-level |  |  |
|  | k6283, k6286, k6287, k6289, k6290, k6291 | A-level |  |  |
|  | k6292 | Degree |  |  |
|  | n4012, n4000 | CSE/none |  |  |
|  | n4003, n4004, n4007, n4015 | Vocational |  |  |
|  | n4001 | O-level |  |  |
|  | n4002, n4005, n4006, n4008, n4009, n4010 | A-level |  |  |
|  | n4011 | Degree |  |  |
| **Age at menarche** | | | | |
|  | d010a, n1120, r2080 | How old were you when your periods first started | Age (years) | Recode:  0: 8 – 11 years  1: 12 – 14 years  2: 15 or older  Missing responses from Questionnaire D were replaced from Questionnaire N and Questionnaire R |
| **Material hardship** | | | | |
| ALSPAC hardship items |  | **How difficult at the moment do you find it to afford these items:** | |  |
|  | c520 , f800 ,g835, h730, k6200, m5170 ,t1360 | a) food | 1: Very difficult  2: Fairly difficult  3: Slightly difficult  4: Not difficult  5: Paid directly by social security | Recode responses of 5 (Paid directly by social security) to 4  Material hardship = 20 – summed scored of 5 questions.  (Higher scores = more material hardship) |
|  | c521, f801, g836, h731, k6201, m5171, t1361 | b) clothing |  |  |
|  | c522, f802, g837, h732, k6202, m5172, t1362 | c) heating |  |  |
|  | c523, f803, g838, h733, k6203, m5173, t1363 | d) rent or mortgage |  |  |
|  | c524, f804, g839, h734, k6204, m5174, t1364 | e) things you need for your children |  |  |
| **Social support** | | | | |
| ALSPAC social support items | d790, e600, f910, g216, k8020, l7020, p4020, s6020 | a) Mother feels she has no-one to share feelings with | 1: Exactly feel  2: Often feel  3: Sometimes feel  4: Never feel | (a), (e): (1=0)(2=1)(3=2)(4=3)  (b), (c), (d), (f), (g), (h), (i), (j): (1=3)(2=2)(3=1)(4=0)  Variable scores then summed for a maximum of 30 (higher score = more social support)  Mode imputation: missing values were put to the mode for that variable. |
|  | d791, e601, f911, g217, k8021, l7021, p4021, s6021 | b) Mother feels her partner provides the emotional support she needs |  |  |
|  | d792, e602, f912, g218, k8022, l7022, p4022, s6022 | c) Mother can share experiences with other mothers |  |  |
|  | d793, e603, f913, g219, k8023, l7023, p4023, s6023 | d) Mother feels her neighbours would help in moments of difficulty |  |  |
|  | d794, e604, f914, g220, k8024, l7024, p4024, s6024 | e) Mother is worried that partner might leave |  |  |
|  | d795, e605, f915, g221, k8025, l7025, p4025, s6025 | f) Mother always has someone to share happiness about child |  |  |
|  | d796, e606, f916, g222, k8026, l7026, p4026, s6026 | g) Partner will take over from mother if she is tired |  |  |
|  | d797, e607, f917, g223, k8027, l7027, p4027, s6027 | h) Mother’s family would help in financial difficulty |  |  |
|  | d798, e608, f918, g224, k8028, l7028, p4028, s6028 | i) Mother’s friends would help in financial difficulty |  |  |
|  | d799, e609, f919, g225, k8029, l7029, p4029, s6029 | j) Mother feels if all fails state would support her financially |  |  |
| **Smoking status** | | | | |
|  | b650, n5000, n5002, n5003, n5004, n5005, r6010, r6012, r6013, r6014, r6015, t5560, MA8010, MA8070 | Ever smoked | Yes/No | Recoded into Never, Ever & Current smoker.  Never and Ever smoker responses were sense checked with historical responses. |
|  | f621, g820, h720, j735, k6180, l5050, l5051, m5160, n5010, r6020, s1300, s1301, t5526, V5526, MA8030, MA8060 | No smoked daily at PRES | None, or daily cigarette brackets (e.g. 1–4 to 30+) |  |
|  | s1302, s1303, t5520, t5521, V5520, V5521, MA8080, n5008, r6018, MA8040 | Mother currently smokes | Yes/No |  |
| **Body mass index (BMI)** | | | | |
|  | dw021, m4221, n1145, p1291, s1291, V4400, XB070, MB4540, fm1ms100, fm2ms100, fm3ms100, fm4ms100 | Height | Metres | Average height across all responses was calculated. BMI (kg/m^2^) |
|  | dw002, m4220, n1140, p1290, s1290, V4410, XB080, MB4580, fm1ms110, fm2ms110, fm3ms110, fm4ms110 | Weight | kg |  |
| **Alcohol intake** | | | | |
|  | f626, g825, h724, k6191, t5500, V5500, MA8220 | How many days in the past month did you have the equivalent of 2 pints of beer, 4 glasses of wine or 4 pub measures of spirit? | 1: Every day  2: more than 10 days  3: 5-10 days  4: 3-4 days  5: 1-2 days  6: none | Recode (4/6 = 0) (3=1) (1/2 =2)  0: Never or ≤4 times/month  1: 2–3 times/week  2: ≥ 4 times/week |
| **History of depression** | | | | |
|  | d171, n1061, r2021 | Had severe depression | 1: Yes, recently  2: Yes, in past  3: No, never | Recode 1/2 = 1; 3 = 0  History of depression was defined as either a report of severe depression or an EPDS ≥13 during the reproductive period.  Reproductive period defined using STRAW stages. |
| **History of antidepressant use** | | | | |
|  | See supplementary Table 2 for all variables |  |  | Recode  (1 = 1) and (2 = 1) and (3 = 1)  (4 = 0)  1: Antidepressant use  0: No antidepressant use  History of antidepressant use was defined as any report of antidepressant use in the reproductive period.  Reproductive period defined using STRAW stages. |

eTable 4. Comparison of PRS for age at natural menopause constructed various P-value thresholds.

| ***P-*value threshold for selecting SNPs** | **Number of SNPs included in the PRS** | **R^2^** | **β** | **SE** | ***P*** |
| --- | --- | --- | --- | --- | --- |
| 5x10^-08^ | 395 | 0.079 | 1.129 | 0.095 | 5.3 x10^-31^ |
| 1x10^-06^ | 548 | 0.086 | 1.183 | 0.095 | 5.7 x10^-34^ |
| 1x10^-04^ | 1335 | 0.103 | 1.299 | 0.095 | 1.6 x10^-40^ |
| 0.001 | 3358 | 0.117 | 1.361 | 0.093 | 4.4 x10^-46^ |
| 0.01 | 11827 | 0.117 | 1.333 | 0.091 | 4.4 x10^-46^ |
| 0.05 | 31515 | 0.118 | 1.289 | 0.087 | 9.4 x10^-47^ |
| **0.1** | **47699** | **0.130** | **1.317** | **0.084** | **2.1 x10^-51^** |
| 0.2 | 71404 | 0.132 | 1.309 | 0.083 | 1.9 x10^-52^ |
| 0.3 | 88860 | 0.133 | 1.303 | 0.082 | 1.3 x10^-52^ |
| 0.4 | 102473 | 0.136 | 1.313 | 0.082 | 6.4 x10^-54^ |
| 0.5 | 113438 | 0.135 | 1.307 | 0.082 | 1.3 x10^-53^ |
| 1 | 143561 | 0.137 | 1.316 | 0.082 | 2.8 x10^-54^ |

R^2^ Proportion of variance in age at natural menopause observed phenotype explained by PRS

β Association per SD increase in PRS with age at natural menopause observed phenotype

SE Standard Error

*P* *P*-value for the association per SD increase in PRS with age at natural menopause observed phenotype

eTable 5. Association between PRS_ANM_ at various p-value thresholds and depression during the perimenopausal or postmenopausal period.

| ***P-*value threshold for selecting SNPs** | **Perimenopause** | | | **Postmenopause** | | |
| --- | --- | --- | --- | --- | --- | --- |
|  | **Odds ratio** | **95% CI** | **P** | **Odds ratio** | **95% CI** | **P** |
| 5x10^-08^ | 1.09 | 0.97 - 1.22 | 0.139 | 0.97 | 0.90 - 1.06 | 0.531 |
| 1x10^-06^ | 1.09 | 0.97 - 1.23 | 0.129 | 0.96 | 0.88 - 1.04 | 0.282 |
| 1x10^-04^ | 1.07 | 0.96 - 1.21 | 0.221 | 0.95 | 0.88 - 1.03 | 0.255 |
| 0.001 | 1.04 | 0.93 - 1.16 | 0.508 | 0.96 | 0.89 - 1.04 | 0.329 |
| 0.01 | 0.95 | 0.85 - 1.06 | 0.378 | 0.93 | 0.86 - 1.00 | 0.063 |
| 0.05 | 1.03 | 0.92 - 1.14 | 0.650 | 0.93 | 0.86 - 1.01 | 0.067 |
| **0.1** | **0.97** | **0.87 - 1.08** | **0.604** | **0.92** | **0.86 - 0.99** | **0.035** |
| 0.2 | 0.96 | 0.86 - 1.07 | 0.457 | 0.91 | 0.85 - 0.98 | 0.018 |
| 0.3 | 0.96 | 0.86 - 1.07 | 0.461 | 0.92 | 0.86 - 1.00 | 0.037 |
| 0.4 | 0.96 | 0.86 - 1.06 | 0.395 | 0.92 | 0.86 - 0.99 | 0.035 |
| 0.5 | 0.96 | 0.86 - 1.06 | 0.391 | 0.92 | 0.85 - 0.99 | 0.024 |
| 1 | 0.95 | 0.85 - 1.05 | 0.316 | 0.92 | 0.85 - 0.99 | 0.028 |

CI: confidence interval; PRS_ANM_: Polygenic risk score for age at natural menopause

eTable 6. Association between age at natural menopause and depression during postmenopause, including all postmenopausal depression assessments.

| **Analysis type** | **Exposure** | **Model** | **Odds ratio** | **95% CI** | ***P*** | **N** |
| --- | --- | --- | --- | --- | --- | --- |
| **Depression during postmenopause** | | | | | | |
| Multivariable regression | Age at menopause (1-yr increase) | Unadjusted | 0.97 | 0.95 – 1.00 | 0.059 | 1,853 |
| Multivariable regression | Age at menopause (1-yr increase) | Adjusted* | 1.00 | 0.97 – 1.03 | 0.972 | 1,853 |
| One-Sample MR | Age at menopause (1-yr increase) | –^+^ | 0.93 | 0.85 – 1.03 | 0.154 | 1,440 |

*Adjusted for age, ethnicity, social class, education, age at menarche, material hardship, social support, smoking status, body mass index and alcohol intake.

^+^Adjusted for the first 10 principal components of ancestry

CI: confidence interval; MR: Mendelian randomization

eTable 7. Association between age at natural menopause and depression during peri- and post-menopause, further adjusting for history of depression and history of antidepressant use as confounders.

| **Analysis type** | **Exposure** | **Odds ratio** | **95% CI** | ***P*** | **N** |
| --- | --- | --- | --- | --- | --- |
| **Depression during perimenopause** | | | | | |
| Multivariable regression | Age at menopause (1-yr increase) | 0.98 | 0.88 – 1.08 | 0.636 | 1,233 |
| **Depression during postmenopause** | | | | | |
| Multivariable regression | Age at menopause (1-yr increase) | 0.98 | 0.92 – 1.04 | 0.511 | 1,587 |

Adjusted for age, ethnicity, social class, education, age at menarche, material hardship, social support, smoking status, body mass index, alcohol intake, history of depression and history of antidepressant use.

CI: confidence interval

eTable 8. Association between age at natural menopause and depression during peri- and post-menopause using only an EPDS score of ≥ 13 to indicate probable depression.

| **Analysis type** | **Exposure** |  | **Odds ratio** | **95% CI** | ***P*** | **N** |
| --- | --- | --- | --- | --- | --- | --- |
| **Depression during perimenopause** | | | | | | |
| Multivariable regression | Age at menopause (1-yr increase) | Unadjusted | 1.02 | 0.98 – 1.06 | 0.313 | 1,271 |
| Multivariable regression | Age at menopause (1-yr increase) | Adjusted* | 0.95 | 0.86 – 1.06 | 0.389 | 1,271 |
| Multivariable regression | PRS_ANM_ (1-SD increase) | Adjusted^†^ | 0.97 | 0.87 – 1.08 | 0.632 | 1,990 |
| One-Sample MR | Age at menopause (1-yr increase) | –^+^ | 1.04 | 0.92 – 1.17 | 0.577 | 1,003 |
| **Depression during postmenopause** | | | | | | |
| Multivariable regression | Age at menopause (1-yr increase) | Unadjusted | 0.98 | 0.94, 1.01 | 0.186 | 1,639 |
| Multivariable regression | Age at menopause (1-yr increase) | Adjusted* | 0.99 | 0.93, 1.06 | 0.840 | 1,639 |
| Multivariable regression | PRS_ANM_ (1-SD increase) | Adjusted^†^ | 0.92 | 0.85, 0.99 | 0.029 | 2,767 |
| One-Sample MR | Age at menopause (1-yr increase) | –^+^ | 0.98 | 0.87, 1.10 | 0.706 | 1,258 |

*Adjusted for age, ethnicity, social class, education, age at menarche, material hardship, social support, smoking status, body mass index and alcohol intake.

^†^Adjusted for age and the first 10 principal components of ancestry

^+^Adjusted for the first 10 principal components of ancestry

EPDS: Edinburgh Postnatal Depression Score; CI: confidence interval; MR: Mendelian randomization, PRS_ANM_: Polygenic risk score for age at natural menopause, SD: standard deviation

eTable 9. Comparison of distribution of confounders at baseline between women included and excluded from analyses. Populations compared for those with complete data on all confounders.

| Characteristic |  | Not included in any analyses | Included in at least one analyses | *P* |
| --- | --- | --- | --- | --- |
| N |  | 5,775 | 3,512 |  |
| Ethnicity, N (%) | White | 5,628 (97.5) | 3,475 (98.9) | <0.001 |
|  | Non White | 147 ( 2.5) | 37 ( 1.1) |  |
| Social class, N (%) | I (highest non-manual) | 130 ( 2.3) | 193 ( 5.5) | <0.001 |
|  | II (non-manual) | 1,131 (19.6) | 1,122 (31.9) |  |
|  | III (non-manual) | 1,431 (24.8) | 970 (27.6) |  |
|  | IV (manual) | 1,798 (31.1) | 783 (22.3) |  |
|  | V (manual) | 980 (17.0) | 364 (10.4) |  |
|  | VI (lowest manual) | 305 ( 5.3) | 80 ( 2.3) |  |
| Age at menarche, N (%) | 8 -11 years | 1,172 (20.3) | 602 (17.1) | <0.001 |
|  | 12-14 years | 3,867 (67.0) | 2,472 (70.4) |  |
|  | 15+ years | 736 (12.7) | 438 (12.5) |  |
| Education, N (%) | CSE/vocational degree | 1,282 (22.2) | 324 ( 9.2) | <0.001 |
|  | O-level | 2,375 (41.1) | 1,113 (31.7) |  |
|  | A-level/University degree | 2,118 (36.7) | 2,075 (59.1) |  |
| Alcohol intake, N (%) | Never or less than 4 times a month | 4,980 (86.2) | 3,015 (85.8) | 0.362 |
|  | 2 to 3 times a week | 522 ( 9.0) | 344 ( 9.8) |  |
|  | 4 or more times a week | 273 ( 4.7) | 153 ( 4.4) |  |
| Smoking status, N (%) | Never | 2,752 (47.7) | 2,030 (57.8) | <0.001 |
|  | Ever | 1,551 (26.9) | 989 (28.2) |  |
|  | Current | 1,472 (25.5) | 493 (14.0) |  |
| Material hardship (mean (SD)) | | 3.19 (3.63) | 2.34 (3.10) | <0.001 |
| Social support (mean (SD)) | | 20.20 (5.45) | 20.84 (5.13) | <0.001 |
| Body mass index (mean (SD)) | | 23.35 (4.15) | 22.81 (3.58) | <0.001 |

eTable 10. Proportion of observations with missing data for each confounder.

| **Confounder** | **Proportion of observations with missing value (%)** |
| --- | --- |
| Ethnicity | 1.72 |
| Social class | 7.66 |
| Education | 1.48 |
| Age at menarche | 3.83 |
| Material hardship | 2.06 |
| Social support | 0.29 |
| Smoking status | 0.00 |
| Body mass index | 0.78 |
| Alcohol intake | 0.58 |

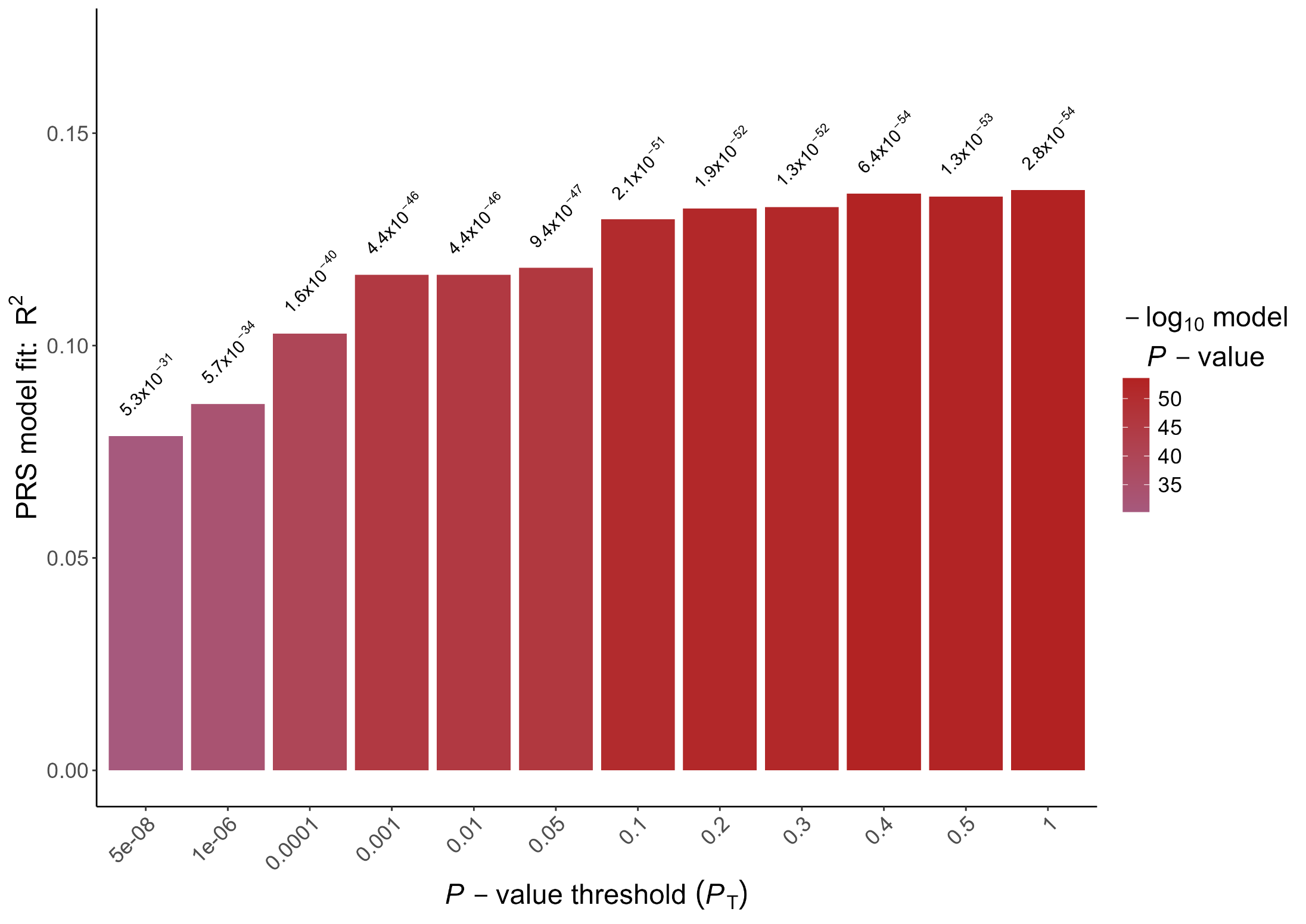
eFigure 1. Variance explained (R^2^) in age at natural menopause by polygenic risk scores (PRS) across *P*-value thresholds.
